# Investigating opportunities to prevent Type 2 Diabetes Mellitus after Gestational Diabetes Mellitus: the DIVINE-NSW study

**DOI:** 10.1101/2022.11.27.22282793

**Authors:** Vivian YJ Lee, Amanda Beech, Angela Makris, Clare Arnott, Janani Shanthosh, Katherine Donges, Anushka Patel, Amanda Henry

## Abstract

**Background:** Gestational Diabetes Mellitus (GDM), once thought to be fully reversed after pregnancy, is now a firmly established independent risk factor for the subsequent development of Type 2 Diabetes Mellitus (T2DM), cardiovascular disease and other chronic conditions. This provides a strong rationale to identify preventive strategies in women with prior GDM, including intervention soon after childbirth. Currently, preventive strategies are mostly focused on modifying lifestyle, with an emphasis on diet and physical activity. However, evidence for the effectiveness of implementing and sustaining changes in behaviour through lifestyle programs is limited, and only a small proportion of women in Australia are thought to engage in lifestyle modification programs. Consideration of additional approaches, including pharmacotherapy, is therefore warranted. The current study aims to 1) measure the prevalence and identify the predictors (up to 4 years post-partum) of persisting dysglycaemia among a diverse population of urban Australian women with recent GDM, 2) understand women’s views and views of their healthcare providers on long-term risks of T2DM and barriers and facilitators to engaging in screening and preventive strategies (including pharmacotherapy) to mitigate these risks, and 3) examine the feasibility of a randomised controlled trial of preventive drug therapies in this population.

**Methods:** This is a retrospective cohort study with a qualitative sub-study. We will identify GDM-affected women who gave birth between January 2018 and December 2021 in at least three Sydney Hospitals (Liverpool Hospital, Royal Hospital for Women and St George Hospital) and invite them to participate in the study. Eligible participants will complete an online questionnaire and an oral glucose tolerance test to assess their current glycaemic status if they have not done so within 12 months of consent and are not currently pregnant. A subset of participants will be invited to participate in an interview to understand their perspectives of GDM, long-term risks and willingness to take preventive medications (including willingness to participate in trials of preventive medicines). Interviews with healthcare providers will also be conducted to understand their views of long-term diabetes risk, screening, and preventive strategies for women following GDM.

**Discussion:** This study will help understand post-GDM care gaps and outcomes currently in Australia, as well as inform the design and conduct of future trials of preventive drug therapies in this population.

**ANZCT Registration:** ACTRN12621001618842

## Introduction

### Background and rationale

The burden of Type 2 Diabetes Mellitus (T2DM) has increased exponentially in Australia and globally in recent decades.[1] Gestational Diabetes Mellitus (GDM), once thought to be fully reversed after pregnancy, is now a firmly established independent risk factor for the development of T2DM, with the overall risk reported to be as high as 10-fold in women with previous GDM.[2] In Australia, GDM prevalence has increased from 5% to 15% between 2000-2017, with major risk determinants including age, ethnicity, and maternal weight.[3] For example, women of Asian ethnicity are twice as likely to develop GDM than those of Caucasian ethnicity, rising to a three-fold increased risk once aged over 30 years.[4] Certain ethnicities are also found to have higher conversion rates to T2DM, including a four-fold greater risk of progression in Aboriginal and Torres Strait Islander women compared to non-Indigenous women.[5] Concerningly, conversion to T2DM following GDM often occurs in the early years after pregnancy. Persisting dysglycaemia is found in 40% of women with GDM within 7 months post-partum in South Asia,[6] and in 72% of women within 5 years in India.[7] Consequently, 25% of women were found to have T2DM within 2 years following GDM, which increased to 50% in 2-4 years.[7] Therefore, there is a need to develop preventive strategies, especially in the early years after pregnancy. The importance of this was recently highlighted in the updated UK NICE guidelines where diabetes prevention is recommended for all women with a previous GDM diagnosis.[8]

Lifestyle interventions, when accessible and engaged with, can be effective in reducing the risk of postpartum diabetes so are generally recommended to be provided to those following GDM.[9, 10] However, the early months and years after pregnancy present a unique challenge to women who often have a key caregiver role, relegating personal health concerns to a lower priority.[11] New mothers also report a lack of time and energy, making lifestyle modification towards a more healthy diet and regular physical activity difficult.[11] This highlights the challenges in achieving behaviour modifications, especially in this population.[6] In addition to lifestyle modifications, currently proposed preventive strategy include pharmacological interventions.[12] Studies have found the thiazolidinedione class of drugs, incretin-based agents, and combined pills (sitagliptin and metformin) [12] to be beneficial, with the strongest evidence associated with metformin.[13-15] This is reflected in the guidelines by the American Diabetes Association (2017) suggesting the use of metformin for prevention of T2DM in those with persisting dysglycaemia, Body Mass Index (BMI) >35 kg/m^2^, age over 60 years and women with prior GDM.[16] However, the use of preventive medications to prevent T2DM in these populations is not common.

Given such high levels of individual and population-level health risk, and the uncertainty of engagement with and ability to implement effective lifestyle interventions at this life stage, there is a compelling need to investigate low-cost and scalable pharmacological preventive approaches. This should ideally be conducted in addition to lifestyle interventions to improve efficacy.[10] While metformin has shown strong evidence for use in reducing the risk of T2DM,[13-15] especially in those at highest risk, including in women with a history of GDM, no definitive evidence currently exists with one feasibility study currently underway in the UK.[17] We propose developing a complementary trial in Australia; this requires critical preliminary data to ensure a robust study design and conduct. This study will build the foundations for future trials in Australia by identifying the prevalence and predictors of T2DM in women who had GDM and examining the feasibility and acceptability of a pharmacological intervention to prevent the development of T2DM in this population.

### Objectives

1. To measure the prevalence and identify the predictors (up to 4 years post-partum) of persisting dysglycaemia among a diverse population of urban Australian women with recent GDM.
2. To identify women’s views and views of their healthcare providers on long-term risks of T2DM and barriers and facilitators to engage in screening and preventive strategies to mitigate these risks.
3. To examine the feasibility of a randomised controlled trial of preventive drug therapies in this population.

### Trial Design

Retrospective cohort study and qualitative sub-study.

## Methods: Participants, interventions, and outcomes

### Study setting

Potential participants will be identified through at least three public hospitals in metropolitan Sydney, Australia (Liverpool Hospital, Royal Hospital for Women and St George Hospital). These hospitals collectively serve a diverse sociodemographic patient population with approximately 11,000 births per year.

### Eligibility criteria

#### Retrospective cohort study

Individuals who are 16 years or older and gave birth following a GDM affected pregnancy between January 2018 and December 2021 will be eligible to participate. Eligible participants are those able to provide written informed consent and are fluent in English, Arabic, Mandarin, Vietnamese, Bangla, or Hindi (the most common languages spoken by the patients in the catchments of the study hospitals). Exclusion criteria include diagnosed diabetes prior to pregnancy and any known inability to provide informed consent to participate due to severe active mental health issues or major developmental disability.

#### Qualitative study

Maximum variation sampling will be used to identify study participants for inclusion in the in-depth interviews. Key dimensions of variation for sampling include age, severity of GDM, perception of risk of developing T2DM, highest level of education, socioeconomic status, and cultural and language background.

Eligible healthcare workers will be obstetricians and endocrinologists employed by one of the three participating hospitals and general practitioners working in hospital catchment areas (identified through shared care arrangements). Eligibility is based on experience in managing patients with GDM. Maximum variation sampling will also be used to identify healthcare providers, with a focus on variation being specialisation, gender, and the number of years post-specialisation.

### Informed consent

Potential participants will be provided (electronically or by mail) a Participant Information Sheet in the language of their choice (English, Arabic, Mandarin, Vietnamese, Bangla, or Hindi). Consent (eConsent or written, depending on the woman’s preference) will be obtained with the availability of telephone contact with study staff to clarify any concerns or questions.

### Study procedures

#### Retrospective cohort study

Eligible and consented participants will be asked to complete an online questionnaire (Appendix 1) exploring views on long-term risks of developing T2DM, and barriers and facilitators to engaging in screening and preventive strategies to mitigate these risks. A yearly oral glucose tolerance test (OGTT) is clinically recommended in those with a history of GDM. If participants have not had one within the previous 12 months of consenting to the study, they will be asked to complete an 75g 2-hour OGTT. Those individuals will be sent a pathology request form by mail or electronically. Participants who are currently pregnant will not be asked to complete an OGTT (Table 1).

**Table 1.** Summary of the criteria for an OGTT.

| If you fall into this category... | This is what we will ask of you |
| --- | --- |
| You HAVE completed an OGTT in the last 12 months regardless of whether or not you are currently pregnant. | We will ask you for a copy of the results or for your permission to access the results from your healthcare provider. |
| You have <u>NOT</u> completed an OGTT in the last 12 months and are currently <u>NOT</u> pregnant. | We will ask you to complete an OGTT at any pathology laboratory most convenient to you (we will send you a request form that can be used at the pathology service of your choice). |
| You have <u>NOT</u> completed an OGTT in the last 12 months and are <u>CURRENTLY</u> pregnant. | We will not ask you to do anything (although please follow your pregnancy care provider's instructions regarding when to do an OGTT for your current pregnancy). |

Additionally, study personnel will access data relating to the participant’s prior GDM-affected pregnancy, GDM treatment and any subsequent pregnancies from the hospital maternity databases and relevant clinics associated with the hospital.

#### Qualitative study

Eligible and consented participants will be asked to participate in an in-depth interview exploring women’s perspectives of GDM, long-term risks and willingness to take preventive medications and to participate in trials of preventive medicines for the study procedures.

**Figure 1.**
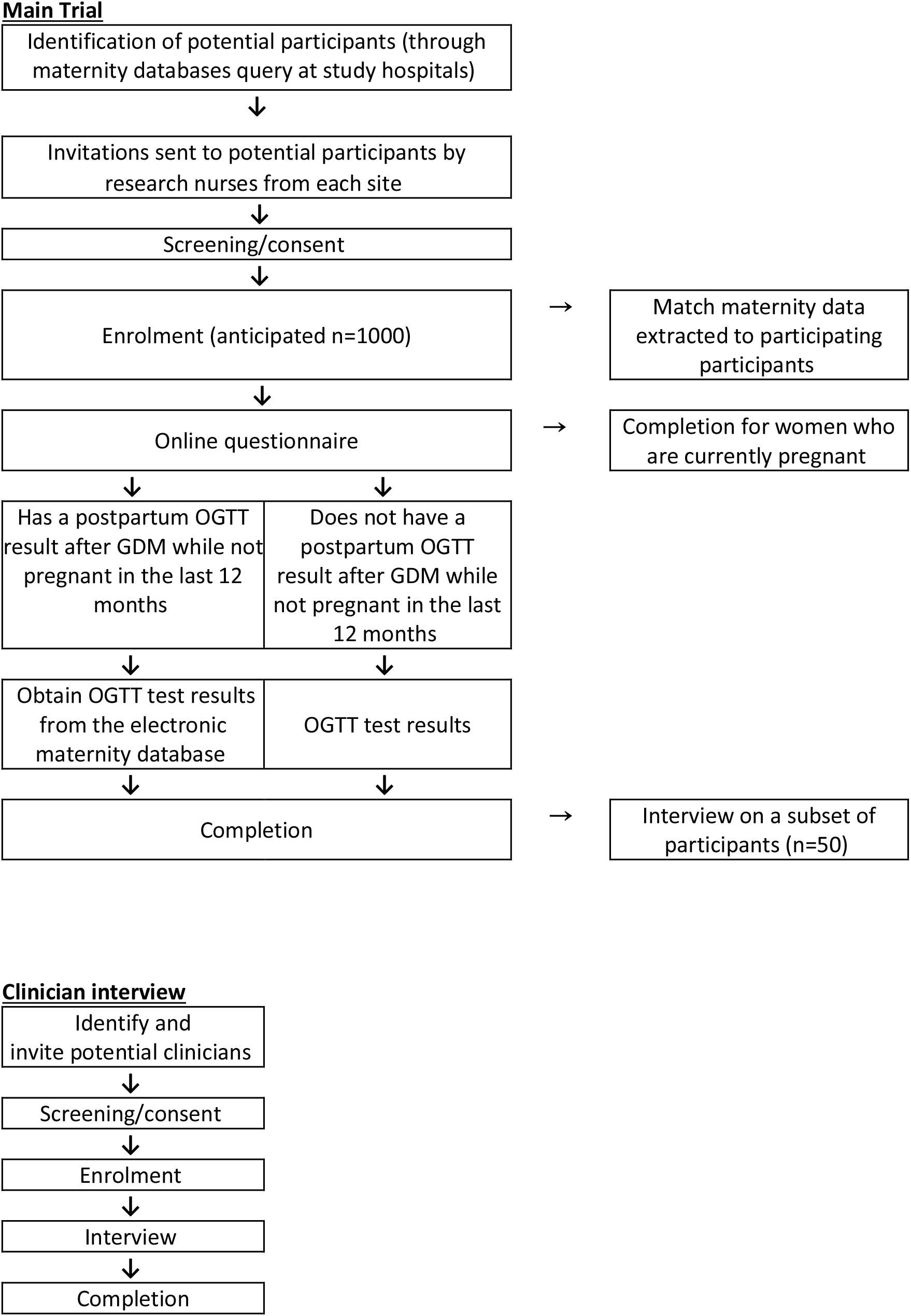
Flowchart of the overall study. Key: OGTT = Oral glucose tolerance test; GDM = gestational diabetes mellitus

### Main Trial

#### Statistical Considerations

Based on existing data from participating hospitals, we expect approximately 4500 eligible women. With an estimate of 20% providing consent,[18] a sample size of approximately 1000 would provide precise overall estimates of post-partum dysglycaemia prevalence. Assuming 60% of participants had a post-partum OGTT within six months of delivery and 10% of those have persisting dysglycaemia, we would expect 80% power (2α=0.05) to detect a relative risk of 1.78 for T2DM development among those with persisting dysglycaemia compared to those with normoglycaemia. This assumes 20% of those with persisting dysglycaemia will have developed T2DM at a median of 18 months follow-up.

Approximately 50 participants across the sites and 10-15 healthcare providers from each hospital and its local GP network will be recruited for the in-depth interviews. The qualitative interviews will continue with thematic analysis of all data contemporaneously against data collection until saturation is achieved. The coding framework development will be informed by Michie’s Behaviour Change Theory (Michie’s Theory).[19] Based on prior studies, we anticipate the proposed sample size will achieve thematic saturation, however this can be increased if required.

Michie’s theory was used to guide the design of our qualitative tools. The theory consolidates key behaviour change frameworks and incorporates a Capability Opportunity Motivation-Behaviour (COM-B) Model. It will provide the ideal framework for generating evidence about the potential acceptability and uptake of pharmacological therapy, and where relevant, its interaction with lifestyle strategies.

Descriptive statistics will be used to report study characteristics, and multivariable modelling used to identify independent predictors of post-partum dysglycaemia, including T2DM. Online questionnaire data will be analysed and reported using descriptive statistics (number and percentage) for closed answer questions and compared between; women who have IGT and those who do not at baseline, who had used medicated GDM to those that did not, ethnicities, and other baseline characteristics (BMI at baseline, number of GDM affected pregnancy) using Chi-squared testing. The open-ended questions will be analysed and reported thematically.

## Discussion

The current study will allow an understanding of the prevalence and predictors of persisting dysglycaemia in a diverse contemporary urban population of Australian women with a recent history of GDM, as well as inform the feasibility and acceptability of a pharmacological intervention to prevent the development of T2DM in this population.

Previously reported prevalence of persisting dysglycaemia in women following GDM affected pregnancy is found to vary, with 23% reported within 12 weeks from delivery in Italy,[20] 40% within 7 months post-partum in South Asia and even up to 48% at a median of 20 months post-partum in India.[21] The most recent estimates of persisting dysglycaemia in the Australian population was reported in 2021, with 22% of First Nation women but no Caucasian women developing T2DM following GDM at a median of 2.5 years follow-up.[22] However, this study had a small sample size (203; 109 First Nations women and 94 Caucasian) and was conducted in the Northern Territory. Therefore, the current study will provide an update on current incidence of GDM in metropolitan Australia.

Currently, studies support the use of certain pharmacological interventions to reduce the risk of T2DM in women with a history of GDM,[12] however the evidence is sparse and has been insufficient to broadly influence guidelines and practice.[14] A better understanding of potential barriers to the use of preventive drug therapy, in addition to the availability of more robust clinical trial data, is needed. A similar study conducted recently in China found women favoured lifestyle changes over pharmacological interventions unless they had family members with diabetes or personally took medications during pregnancy.[23] The qualitative data in the current study are expected to generate new evidence about the potential acceptability and uptake of pharmacological therapy, and where relevant, its interaction with lifestyle strategies. This approach will enable us to investigate, from the perspectives of patients and their healthcare providers, women’s motivations (including unconscious and conscious thoughts and goals) that influence behaviour, for example, engagement with lifestyle strategies and the uptake and adherence to medications. It will also investigate social and physical opportunities available to women to perform these behaviours as well as physical and psychological capabilities (including knowledge, skills, and tools).

## Strengths and Limitations

This study will help to examine current post-GDM care and outcomes in Australia. It will assist in understanding care gaps for women with prior GDM and provide critical information towards the design and conduct of a large-scale trial to test the usefulness and safety of drug therapies to prevent long-term diabetes among women who had GDM. Strengths include having a large sample size which will provide precise estimate of incidence and power to identify independent predictors of persisting dysglycaemia. This should also allow us to assess consecutive recruitment onto potential future large-scale trials within a diverse catchment population. Some limitations include potential self-selection bias in participating in the study, which is not yet known, and the findings may have limited generalisability.

## Trial Status

Current protocol version 5.0 dated 08/03/2022. Approximate recruitment and data collection between June 2022-June 2023.

## Data Availability

All data produced in the present work are contained in the manuscript

## Abbreviations

GDM: Gestational Diabetes
T2DM: Type 2 Diabetes Mellitus
OGTT: Oral glucose tolerance test

## Declaration

### Authors’ contributions

All the included authors are a part of the steering committee. AH and AP are the PIs and co-chair the steering committee. AH and VL will obtain appropriate ethics and governance approvals and work with all research staff throughout the study duration. AH, AB and AM will oversee research staff at each of the three clinical sites. JS will be responsible for the qualitative aspect of the study and conduct interviews. KD is the consumer and community involvement representative. The team will work together for data interpretation and dissemination of the results.

### Availability of data and material

The final dataset will be kept in a secure server at The George Institute for Global Health, and only relevant research staff will have access to the data.

### Ethics approval and consent to participate

Ethical approval has been obtained by the South Eastern Sydney Local Health District HREC (2022/ETH00276).

## Appendix 1

### Section 1: About you

These first few questions are to find out about you and your background.

1. What is your height? ____________cm
2. What is your weight? ____________kg
3. Please choose the highest educational qualification you have received.
  □ Never attended school
  □ Year 9 or below
  □ Secondary school (Year 10-12 or equivalent)
  □ Trade Certificate/Diploma
  □ University Degree
  □ Prefer not to answer

### Section 2: Health and Diabetes

4. Are you currently diagnosed with any of the following non-diabetes conditions? (Please select all that apply).
  □ Hypertension (High blood pressure)
  □ Hyperlipidaemia (High blood cholesterol)
  □ Polycystic Ovarian Syndrome (“PCOS”)
  □ Autoimmune disease (e.g., rheumatoid arthritis, lupus)
  □ Other major health condition, please state ……………………………………………
5. How many times have you given birth (either preterm or full-term)?
  □ Once
  □ Twice
  □ Three times
  □ Four or more times
6. How many pregnancies have you had affected by gestational diabetes?
  □ One
  □ Two
  □ Three
  □ Four or more
7. Do you have a family history of diabetes?
  □ Yes
  □ No (skip to Question 8)
  □ Not sure (skip to Question 8)
    a. If yes, which type? (Choose all that apply)
      □ Diabetes - type 1 - (usually childhood/early adult onset: require insulin to stay alive)
      □ Diabetes - type 2 - (usually adult onset, do not require insulin to stay alive although insulin may be part of treatment)
      □ Diabetes - type unknown
      □ High sugar levels (HSL)/ “pre-diabetes” /insulin resistance
      □ Not sure
  8. Are you currently diagnosed with diabetes?
    □ Yes
    □ No, but I have been told I have High sugar levels (HSL)/ “pre-diabetes” / insulin resistance (go to Q9)
    □ No (skip to Q9)
      a. If yes, which type?
        □ Diabetes - type 1 - (insulin-dependent)
        □ Diabetes - type 2 - (non-insulin-dependent)
        □ Diabetes - type unknown
        □ Not sure
      b. If yes, how soon after your <u>first</u> GDM-affected pregnancy were you diagnosed?
        □ Six months or less afterwards
        □ More than 6 months and up to12 months afterwards
        □ More than 1 year and up to-2 years afterwards
        □ More than 2 years afterwards
        □ Not sure/can’t remember
      c. If yes, are you on any medication(s)?
        □ Yes
        □ No
      d. If yes, please list all medication(s) you take, including dosage: (skip to Question 13)
    ____________________________________________________________________

**<u>Questions only applicable to those that have answered ‘No’ to Question 8</u>**.

9. How would you rate your risk of developing diabetes in the next 5 years?

10. How concerned are you about developing diabetes?

|  | 1 | 2 | 3 | 4 | 5 | 6 | 7 | 8 | 9 | 10 |  |
| --- | --- | --- | --- | --- | --- | --- | --- | --- | --- | --- | --- |
| No risk | ☐ | ☐ | ☐ | ☐ | ☐ | ☐ | ☐ | ☐ | ☐ | ☐ | Very high risk |

11. How important is it for you to prevent or delay the onset of diabetes?

10. How concerned are you about developing diabetes?
|  | 1 | 2 | 3 | 4 | 5 | 6 | 7 | 8 | 9 | 10 |  |
| --- | --- | --- | --- | --- | --- | --- | --- | --- | --- | --- | --- |
| Not concerned | ☐ | ☐ | ☐ | ☐ | ☐ | ☐ | ☐ | ☐ | ☐ | ☐ | Very concerned |

12. How much effort are you putting in to reduce your risk of diabetes?

11. How important is it for you to prevent or delay the onset of diabetes?
|  | 1 | 2 | 3 | 4 | 5 | 6 | 7 | 8 | 9 | 10 |  |
| --- | --- | --- | --- | --- | --- | --- | --- | --- | --- | --- | --- |
| Not important | ☐ | ☐ | ☐ | ☐ | ☐ | ☐ | ☐ | ☐ | ☐ | ☐ | Very Important |

12. How much effort are you putting in to reduce your risk of diabetes?
|  | 1 | 2 | 3 | 4 | 5 | 6 | 7 | 8 | 9 | 10 |  |
| --- | --- | --- | --- | --- | --- | --- | --- | --- | --- | --- | --- |
| No effort | ☐ | ☐ | ☐ | ☐ | ☐ | ☐ | ☐ | ☐ | ☐ | ☐ | Maximum effort |

**<u>Questions applicable to all</u>**

### Section 3: Lifestyle since your GDM pregnancy

(if more than one pregnancy affected by GDM, please give answers regarding your <u>most recent</u> GDM-affected pregnancy).

13. Did you breastfeed (including expressed breastmilk) following your GDM affected pregnancy?
  □ Yes, I am currently breastfeeding
  □ Yes
  □ No (Skip to Question 14)
    a. If yes, how long did you do *any* breastfeeding (or express breastmilk) for/how long have you currently been breastfeeding for (or expressing breastmilk) after giving birth? (This includes exclusive breastfeeding and mixed breast/bottle feeding).
      □ Less than 2 weeks
      □ Between 2 weeks and 1 month
      □ More than 1 month and up to 2 months
      □ More than 2 months and up to 4 months
      □ More than 4 months and up to 6 months
      □ More than 6 months and up to 9 months
      □ More than 9 months and up to12 months
      □ More than 12 months
14. Have you made any deliberate lifestyle (non-medication) changes since experiencing gestational diabetes (choose all that apply)?
  □ Dietary changes
  □ Physical activity changes
  □ Both dietary and physical activity changes
  □ Other (Please specify) ______________________________________
  □ No changes
    a. If no changes, please let us know the reasons (choose all that apply):
      □ I was already eating a healthy diet
      □ I was already physically active
      □ I was already eating a healthy diet and physically active
      □ No time/too much effort to make these changes
      □ No one advised me to make any lifestyle changes
      □ No one gave me specific advice to help me make lifestyle changes
      □ Too expensive to make lifestyle changes
      □ My family/living circumstances make it hard to make any changes
      □ I am not interested in making any changes
      □ Other (Please specify) ____________________________________
15. How fit do you consider yourself to be?
  □ Very unfit
  □ Unfit
  □ Average
  □ Fit
  □ Very fit
16. We would like to find out about your physical activity levels at certain times. **DEFINITION OF ACTIVITY LEVELS** <u>Not active</u>
  - No planned activity outside of everyday home/work-related activities.
  <u>Slightly active</u>
  - Low-intensity activity (can speak comfortably during exercise) outside for **<u>less than</u>** 150 minutes per week (e.g., walks).
  <u>Active</u>
  - Planned moderate-intensity (unable to complete a sentence without a break during exercise) exercise of **<u>at least</u>** 150 minutes per week.
  **OR**
  - **<u>At least</u>** 60 minutes of vigorous-intensity (can only say a few words at a time) exercise per week.
  Very active
  - Planned moderate-intensity (unable to complete a sentence without a break) exercise of **<u>more than</u>** 150 minutes per week.
  **OR**
  - **<u>More than</u>** 60 minutes of vigorous-intensity (can only say a few words at a time) exercise per week.
  Please tick one of the following.

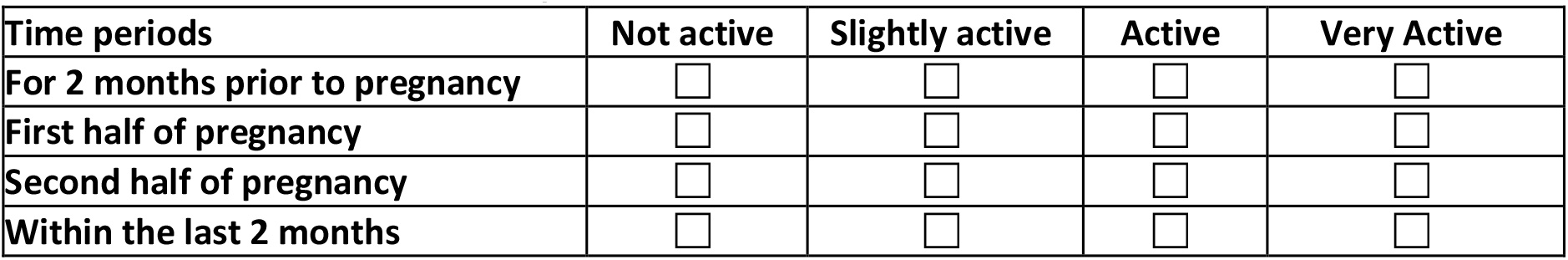

**<u>Question applicable to only those that have answered ‘no’ to question 8</u>**.

### Section 4: Perspectives on taking preventative medications

17. How likely would you take on lifestyle changes (diet, exercise) to prevent you getting diabetes?

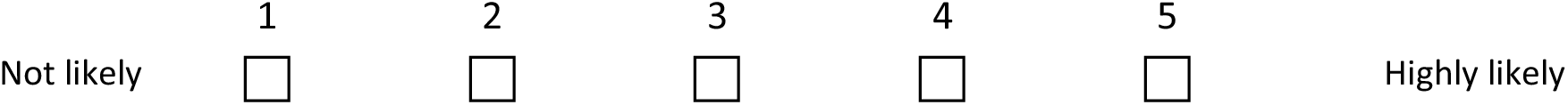
18. How likely would you take preventative medication proven safe and effective to prevent you getting diabetes?

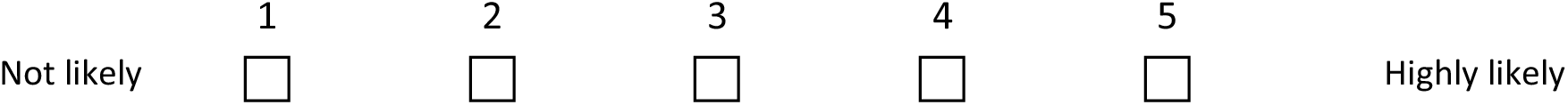
19. If you would consider taking medication to prevent you getting diabetes, how long would you be prepared to take medication for?
  □ Up to six months
  □ More than six months and up to a year
  □ More than 1 year and up to2 years
  □ More than 2 years but not forever
  □ As long as needed to prevent me getting diabetes
  □ I do not want to take any medications
20. If you would consider taking medication to prevent you getting diabetes, what types of medication would you be prepared to take? (Please tick all that apply).
  □ Tablets or capsules (oral medications)
  □ Needles/injected medication that you give to yourself
  □ Injected medication given to you by a doctor or nurse
  □ Not sure
  □ I do not want to take any medications
21. If you are concerned about taking medications to prevent the onset of diabetes, what are your concerns? (Choose all that apply).
  □ I am worried about the side effects of medication
  □ I am worried about medication safety if I have another baby
  □ I am worried about medication safety while breastfeeding
  □ I am worried about the cost
  □ I do not like taking medications
  □ I do not trust medications
  □ I am not concerned about getting diabetes
  □ I am concerned about getting diabetes but not enough to take medication
  □ Other, please specify: _____________________________________________
  □ I am not concerned about taking medications/I am OK with taking medications
22. Would you be interested in being involved in research studies of medicines to prevent diabetes after GDM?
  □ No
  □ Not sure
  □ Yes

18. How likely would you take preventative medication proven safe and effective to prevent you getting diabetes?
|  |  |  |  |  |  |  |
| --- | --- | --- | --- | --- | --- | --- |
|  | 1 | 2 | 3 | 4 | 5 |  |
| Not likely | ☐ | ☐ | ☐ | ☐ | ☐ | Highly likely |

